# Circulating cell-free DNA methylation profiles as noninvasive multiple sclerosis biomarkers: A proof-of-concept study

**DOI:** 10.1101/2025.02.14.25322180

**Authors:** Hailu Fu, Kevin Huang, Wen Zhu, Lili Zhang, Ravi Bandaru, Shruthi Venkatesh, Elizabeth Walker, Li Wang, Yaping Liu, Zongqi Xia

## Abstract

In chronic neurodegenerative diseases such as multiple sclerosis (MS), there is a critical unmet need for cost-effective non-invasive blood biomarkers to concurrently classify disease subtypes, monitor disability severity, and predict long-term progression. In this proof-of-concept study, we performed low-coverage whole-genome bisulfite sequencing (WGBS) on 75 plasma cell-free DNA (cfDNA) samples collected from a well-characterized real-world prospective clinic cohort with longitudinal disability outcome measurements. We assessed the clinical utility of cfDNA methylation profile for differentiating MS patients from non-MS controls, classifying MS subtypes, estimating disability severity, and predicting disease trajectories. We identified thousands of differentially methylated CpGs and hundreds of differentially methylated regions (DMRs) that significantly distinguished MS from controls, separated MS subtypes, and stratified disability severity levels. These DMRs were highly enriched in immunologically and neurologically relevant cis-regulatory elements (*e.g.,* active promoters and enhancers) and enriched in motifs associated with neuronal function and T-cell differentiation, underscoring the interplay between the peripheral immune system and the central nervous system in MS. To distinguish MS subtypes and disability severity levels, models using DMRs achieved area-under-the-curve (AUC) values ranging from 0.67 to 0.81 and models using inferred tissue-of-origin patterns from cfDNA methylation achieved AUC 0.70 to 0.82, outperforming historical benchmark neurofilament light chain (NfL) and glial fibrillary acidic protein (GFAP) in the same cohort. Finally, a linear mixed-effects model identified “prognostic regions” where baseline cfDNA methylation levels were associated with subsequent disability progression and could predict the future disability severity (AUC=0.74) within a 3-year evaluation window. Using these prognostic regions, we generated a cfDNA methylation-based progression risk score for each patient and stratified patient groups with differential progression risks. As we generate higher-depth WGBS data and validate in follow-up studies, the present findings suggest the potential clinical utility of circulating cfDNA methylation profiles as promising MS biomarkers for future validation.

## Introduction

Multiple sclerosis (MS) is a chronic autoimmune disease impairing the central nervous system (CNS), causing inflammatory demyelination and progressive neurodegeneration in approximately three million individuals globally.^1,2^ Most individuals develop initially Relapsing-Remitting MS (RRMS), defined by discrete episodes of acute neurological symptoms (*i.e.,* relapse) followed by partial or full recovery (*i.e.,* remission), and subsequently transition to Secondary Progressive MS (SPMS) after decades, while a subpopulation develop Primary Progressive MS (PPMS) from the onset.^3^ Patients with progressive MS (PMS), including both PPMS and SPMS, experience worsening neurological impairment without discrete periods of relapse or remission.^4^

Among the approved disease-modifying therapies (DMTs),^5,6^ all have indications for RRMS or “active” SPMS (*i.e.,* with ongoing relapses). No DMT is yet approved for non-active SPMS (*i.e.,* without relapse), and only one DMT has indication for PPMS as of this study.^7,8^ Early initiation of suitable DMTs improves long-term outcomes, whereas inappropriate DMT selection may cause suboptimal outcomes. Timely MS subtype classification could improve DMT selection, but MS subtyping is retrospective in current practice, potentially delaying appropriate therapeutic guidance. Further, given patient variations in disability progression and treatment response, effective monitoring of disease severity and iterative prognosis could improve clinical care. Magnetic resonance imaging (MRI) is the chief monitoring method in current practice, but has prediction limitations and practical constraints due to access difficulties, patient discomfort, high cost, and insurance restriction.^1,9,10^

Non-invasive blood biomarkers provide accessible and cost-effective alternatives for MS subtype classification, disability monitoring, and disease prognosis.^11^ Even as biomarkers like neurofilament light chain (NfL)^12^ and glial fibrillary acidic protein (GFAP)^13^ enter clinical practice, there are rooms for improvement throughout the clinical contexts of MS care. Although elevated NfL and GFAP levels indicate neuronal and astrocytic injury, respectively, they do not fully capture the multifaceted pathology underlying inflammatory disease activity, progression from RRMS to SPMS, or the steady decline in PMS.^14–16^

Circulating cell-free DNA (cfDNA) from peripheral blood has emerged as a non-invasive, accessible, and cost-effective biomarker for multiple diseases.^17–20^ It is well suited for capturing complex pathogenesis underlying chronic autoimmune diseases such as MS, which involves peripheral immune activation, inflammatory demyelination and CNS neurodegeneration.^21^ In people with MS (pwMS), cfDNA can originate from multiple cell types undergoing apoptosis or necrosis, including T cells, B cells, and monocyte cells of the peripheral immune system as well as oligodendrocytes and neurons of the CNS.^22^ In previous studies, distinct cfDNA methylation patterns at individual genomic regions differentiated pwMS from controls.^23–25^ Genome-wide and tissue-specific studies reported widespread aberrant DNA methylation patterns in the peripheral immune system and CNS of pwMS,^26–32^ which collectively add to the circulating cfDNA pool and justify a strategy of concurrently profiling the peripheral immune system and CNS. Given the cell-type specificity of DNA methylation, cfDNA methylation signatures could indicate the tissues of origin, enabling a detailed analysis of underlying pathological processes.^33–37^

We hypothesized that circulating cfDNA methylation profiles would have clinical utility in subtyping MS, monitoring disability severity, and predicting disease progression and could eventually complement current methods (*e.g.,* MRI, existing blood biomarkers). For instance, a greater proportion of CNS-derived cfDNA might suggest progressive MS subtypes or worse neurodegeneration, whereas elevated peripheral immune cells-derived cfDNA may indicate acute inflammatory disease activity. Clinical implementation of cfDNA methylation profiles could facilitate earlier initiation or discontinuation of subtype-specific DMTs, more frequent monitoring, and iterative prognosis adjustments tailored to the evolving patient profiles.

To demonstrate the clinical utility, it is crucial to examine the extent to which biomarkers perform in relevant clinical contexts using *affordable* methods. As a proof-of-concept, we performed low-coverage (∼1.5×) whole-genome bisulfite sequencing (WGBS) on 75 plasma samples from pwMS and controls in a well-characterized real-world prospective clinic cohort with longitudinal disability outcome measurements. We assessed the genome-wide differences in cfDNA methylation signature and its inferred tissue-of-origin between pwMS and controls, across MS subtypes, and between pwMS having severe vs non-severe disability status. Using cfDNA methylation profiles and machine learning, we differentiated pwMS from controls, across MS subtypes, and between disability severity levels. Finally, we deployed a linear mixed model to identify “prognostic regions” in baseline cfDNA methylation profile associated with subsequent disability progression after confounder adjustment and generated a cfDNA methylation-based progression risk score (MBPRS) using these prognostic regions to stratify patients with differential progression risks.

## Results

### Differentially methylated CpGs and regions separate MS vs controls and across MS subtypes

We collected 2 mL of plasma from 57 pwMS and 18 non-MS controls enrolled in a clinic-based prospective cohort (*i.e.,* PROMOTE) (**Table 1**). Among the MS samples, 12 were obtained within 3 months of an active relapse from RRMS, 27 during stable remission from RRMS, and 18 from PMS (*i.e.,* PPMS or SPMS) (**Fig. 1**, **Supplementary Table 1**). We generated cfDNA WGBS data with a median of ∼56 million paired-end reads per sample. After removing low-quality reads, we retained a median of ∼34 million paired-end reads per sample, covering ∼22 million CpGs at 1.5× per CpG (**Supplementary Table 2**).

**Figure 1.**
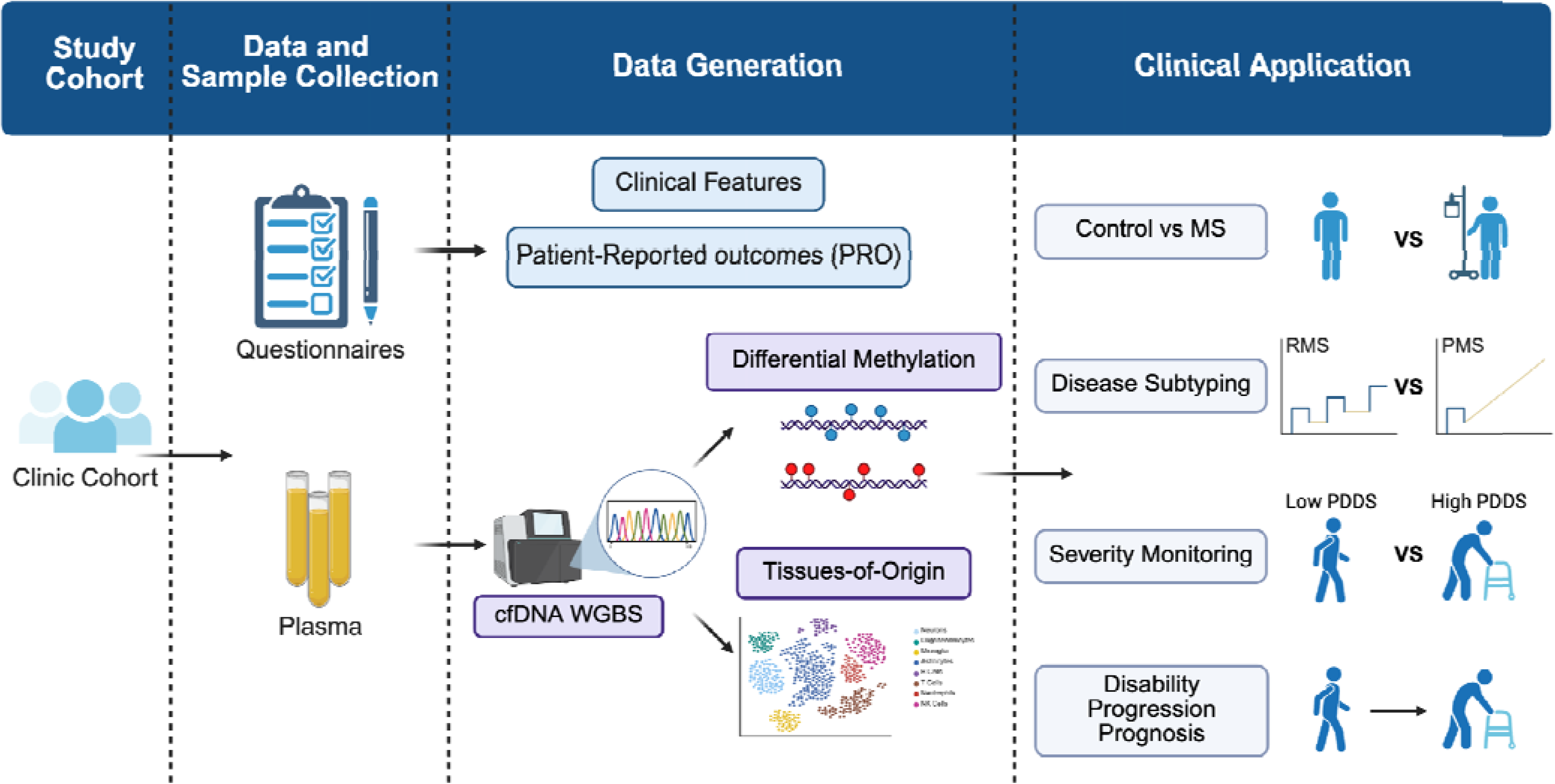
Overview of the study design.

**Table 1.** Participant profile.

|  | Control<br>(N=18) | A-RRMS<br>(N=12) | S-RRMS<br>(N=27) | PMS<br>(N=18) | Total<br>(N=75) | p value |
| --- | --- | --- | --- | --- | --- | --- |
| **Age at Sample Collection**<br>[Y, Mean (SD)] | 57.3 (11.0) | 41.5 (9.5) | 48.3 (11.6) | 61.1 (7.7) | 52.5 (12.3) | 0.053 |
| **Disease Duration**<br>[Y, Mean (SD)] | NA | 14.5 (11.4) | 19.7 (14.2) | 27.5 (12.3) | 21.1 (13.8) |  |
| **Sex [N (%)]** |  |  |  |  |  | 0.017 |
| **Female** | 9 (50.0%) | 7 (58.3%) | 23 (85.2%) | 15 (83.3%) | 54 (72.0%) |  |
| **Male** | 9 (50.0%) | 5 (41.7%) | 4 (14.8%) | 3 (16.7%) | 21 (28.0%) |  |
| **Race [N (%)]** |  |  |  |  |  | 0.522 |
| **Asian** | 0 (0.0%) | 0 (0.0%) | 1 (3.7%) | 0 (0.0%) | 1 (1.3%) |  |
| **Black** | 1 (5.6%) | 3 (25.0%) | 2 (7.4%) | 3 (16.7%) | 9 (12.0%) |  |
| **White** | 17 (94.4%) | 9 (75.0%) | 24 (88.9%) | 15 (83.3%) | 65 (86.7%) |  |
| **Ethnicity [N (%)]** |  |  |  |  |  | NA |
| **Not Hispanic or Latino** | 18 (100.0%) | 12 (100.0%) | 27 (100.0%) | 18 (100.0%) | 75 (100.0%) |  |
| **PDDS [N (%)]** |  |  |  |  |  | <0.001 |
| PDDS ≤1 | 13 (72.2%) | 8 (66.7%) | 11 (40.7%) | 1 (5.6%) | 33 (44.0%) |  |
| **PDDS <4** | 13 (72.2%) | 9 (75.0%) | 17 (63.0%) | 5 (27.8%) | 44 (58.7%) |  |
| PDDS ≥4 | 1 (5.6%) | 3 (25.0%) | 9 (33.3%) | 11 (61.1%) | 24 (32.0%) |  |
| PDDS ≥6 | 1 (5.6%) | 2 (16.7%) | 3 (11.1%) | 7 (38.9%) | 13 (17.3%) |  |

Note: *RRMS*, relapsing-remitting MS; *A-RRMS*, actively relapsing RRMS; *S-RRMS*, stable remission RRMS; *PMS*, progressive MS; *PDDS*, patient determined disease steps (*i.e.*, a patient-reported outcome of disability severity)

Using a beta-binomial regression model (RADMeth^38^), we identified significant DNA methylation changes between pwMS and controls as well as across MS subtypes (**Fig. 2A**). Specifically, we detected 662 hypermethylated and 1,445 hypomethylated CpGs in pwMS compared to controls (FDR<0.1, absolute methylation difference>0.1). Applying the same model and thresholds, we found 487 hypermethylated and 704 hypomethylated CpGs in PMS relative to RRMS. Similarly, we observed 692 hypermethylated and 370 hypomethylated CpGs in active-relapsing RRMS (A-RRMS) compared to stable-remission RRMS (S-RRMS) (**Supplementary Table 3**).

**Figure 2.**
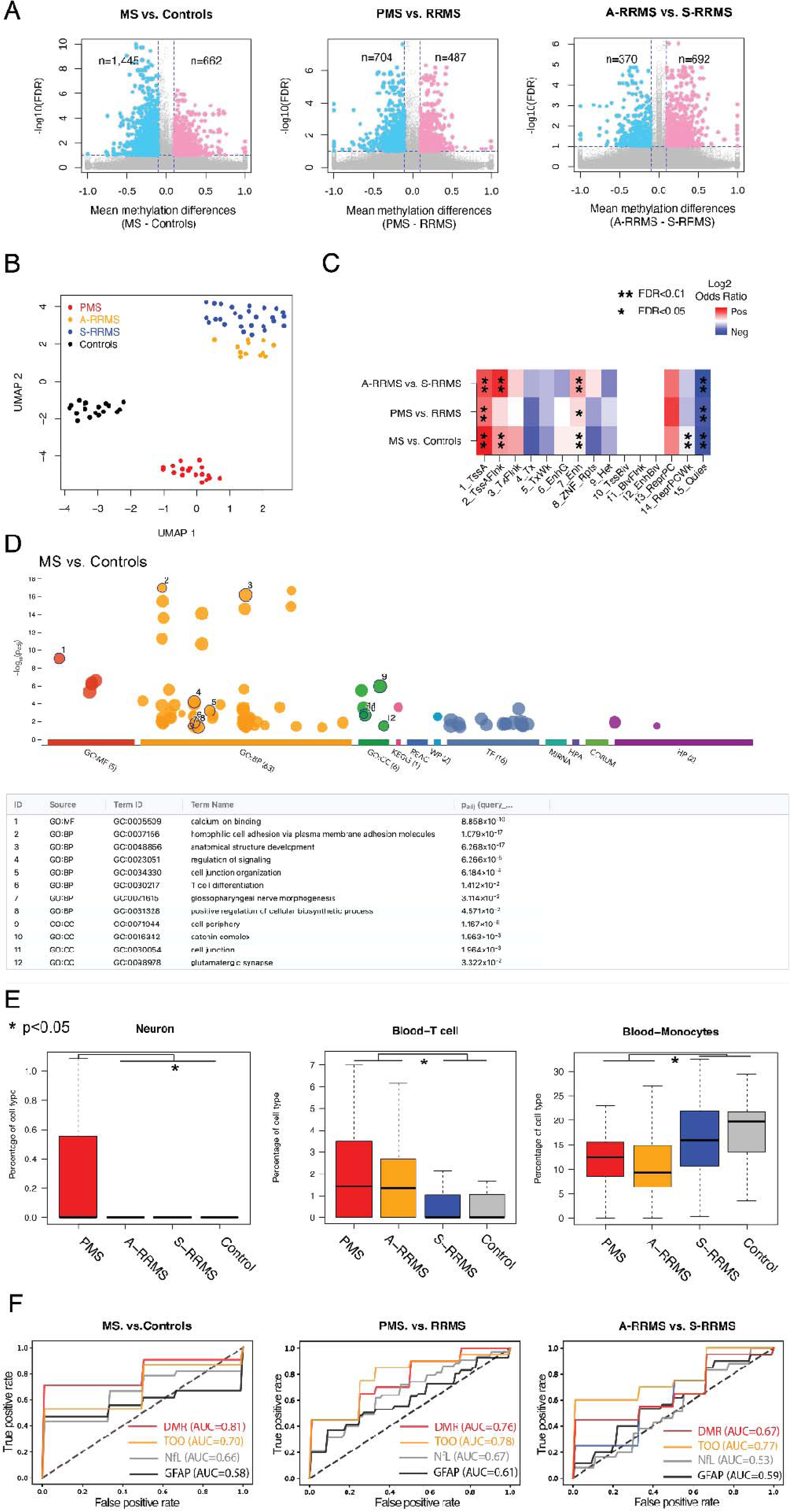
Circulating cfDNA methylation and its inferred tissue-of-origin patterns distinguished MS from controls and among different MS subtypes. **A**. Scatter plots of -log10(FDR) vs mean methylation differences at differentially methylated CpGs between MS and controls (left panel), between progressive MS (PMS: PPMS or SPMS) and relapsing-remitting MS (RRMS) (middle panel), and between actively relapsing RRMS (A-RMSS) and stable remission RRMS (S-RMSS) (right panel). Colors indicate CpGs with FDR < 0.1 and absolute mean methylation difference > 0.1. **B**. Uniform Manifold Approximation and Projection (UMAP) representation of cfDNA methylation levels in DMRs could distinguish pwMS from controls and across MS subtypes. **C**. Enrichment and depletion of DMRs in different chromHMM states (that were previously computed from GM12878 cells in the Epigenome Roadmap project) when comparing MS vs controls or when comparing between MS subtypes. **D**. Gene ontology enrichment analysis of DMRs between MS and non-MS controls with prioritization by g:profile. (See **Supplementary Figure 2** for the gene ontology enrichment analysis of DMRs between PMS and RRMS as well as between A-RMSS and S-RRMS.) **E**. Differences in cell death contribution from brain Neuron (left panel), T cells (middle panel), and Monocytes (right panel) when comparing MS with controls and when comparing across MS subtypes. **F**. Receiver operating characteristic (ROC) curve by DMR only (red), Tissue-of-origin only (TOO, orange), NfL only (grey), and GFAP only (black) to differentiate MS and controls (left panel), PMS and RRMS (middle panel), and A-RRMS and S-RRMS (right panel). NfL and GFAP were measured from a subset of the participants from the same cohort as approximate historical benchmarks.

We next merged adjacent differentially methylated CpGs into differentially methylated regions (DMRs) using RADMeth. This yielded 672 DMRs when comparing pwMS vs controls, 342 DMRs when comparing PMS vs RRMS, and 387 DMRs when comparing A-RRMS vs S-RRMS (**Supplementary Table 4**). Due to the inherent noise in methylation measurements from low-coverage WGBS, which might hinder reliable clinical correlations, we adopted a strategy previously developed for single-cell epigenetic analyses.^39,40^ We performed singular value decomposition (SVD)^41^ on the DNA methylation matrix for DMRs and selected the top components that explained the largest fraction of variability (**Supplementary Fig. 1A**). We then applied Uniform Manifold Approximation and Projection (UMAP)^42^ for dimensional reduction, revealing distinct separations between MS and controls and among the MS subtypes (**Fig. 2B**).

### DMRs across MS subtypes are associated with neuronal and immune-related gene regulatory processes

To assess the gene regulatory potential of these DMRs, we leveraged chromHMM states characterized in immune cells.^43,44^ All identified DMRs showed significant enrichment in active promoter and enhancer states and depletion in quiescent states (**Fig. 2C**, FDR<0.05, Fisher Exact test). DMRs differentiating MS from controls and DMRs distinguishing A-RRMS from S-RRMS were enriched at transcription start site (TSS) flanking states (FDR<0.01, Fisher Exact test), while DMRs differentiating MS and controls further showed significant depletion in weak Polycomb repressive states (FDR<0.01, Fisher Exact test).

In Gene Ontology analysis, these DMRs were highly enriched in both neuronal and immune-related processes (**Fig. 2D, Supplementary Fig. 2, Supplementary Table 5)**. For instance, DMRs distinguishing MS from controls were enriched in T cell differentiation, calcium ion binding, cell adhesion, and anatomical structure development, whereas DMRs separating A-RRMS and S-RRMS were enriched in synaptic signaling. Motif analysis further revealed multiple transcription factor binding sites (TFBSs) significantly overrepresented in all DMRs (**Supplementary Fig. 3, Supplementary Table 5**). For example, ZBTB14, a regulator of monocyte and macrophage development,^45^ was the most enriched TFBS in DMRs, distinguishing between MS and controls. ZNF609, a modulator of myoblast proliferation,^46^ was the most enriched TFBS in DMRs, distinguishing between PMS and RRMS. Finally, ZNF37A, previously implicated in myotonic dystrophy,^47^ was the most enriched TFBS in DMRs, distinguishing A-RRMS from S-RRMS.

### Tissue-of-origin inferred from cfDNA methylation varied significantly across MS subtypes

The tissue-of-origin of cfDNA molecules can be identified through their DNA methylation profiles.^33,35,37^ By deconvoluting cfDNA methylation haplotypes, we observed significantly increased neuronal cell death in PMS compared to other subgroups, and elevated T cell death in both PMS and A-RRMS relative to S-RRMS and controls (**Fig. 2E, Supplementary Fig. 4, Supplementary Table 6,** one-side Mann-Whitney U test, p<0.05). These findings align with previous reports highlighting neuronal cell death in PMS and T cell-mediated pathogenesis in MS.^48^ Interestingly, we also detected significantly lower monocyte cell death and higher megakaryocyte cell death in PMS and A-RRMS, along with reduced erythroid progenitor cell death specifically in the A-RRMS group (**Supplementary Fig. 4**, one-side Mann-Whitney U test, p<0.05). While MS pathogenesis research has investigated T cell–mediated mechanisms, evidence also suggests the involvement of other cell types, including peripheral monocytes^49^ and megakaryocytes.^50^ We interpret these results with caution given the low-coverage WGBS data, which might limit the power to detect tissue-of-origin differences due to the high variability in cfDNA source estimation. Based on simulation analysis, higher coverage (≥8×) WGBS data will be necessary to validate these observations (**Supplementary Fig. 5**).

### cfDNA methylation level and its inferred tissue-of-origin informed MS diagnosis and subtype classification

We examined whether DMR-based cfDNA methylation levels or tissue-of-origin patterns could distinguish MS from controls and among MS subtypes (**Fig. 2F**). Using 10-fold cross-validations, we first identified DMRs and significantly differentiated tissue-of-origin patterns (in CNS and immune cells) within a training set. We then trained machine-learning models on these features and evaluated their performance in the held-out test set. For distinguishing MS from controls, DMRs alone achieved an area under the curve (AUC) of 0.81 (standard deviation [SD] 0.17), while tissue-of-origin features alone yielded AUC 0.70 (SD 0.19). DMRs and tissue-of-origin features outperformed neurofilament light chain (NfL: AUC 0.66) or glial fibrillary acidic protein (GFAP: AUC 0.58) measured in a previous study from the same clinic-based cohort as historical comparison in a subset of the current study.^51^ For distinguishing PMS from RRMS, DMRs achieved AUC 0.76 (SD 0.20) while tissue-of-origin features achieved AUC 0.78 (SD 0.21), outperforming NfL (AUC 0.67) and GFAP (AUC 0.61) in historical comparisons. For distinguishing A-RRMS from S-RRMS, DMRs achieved AUCs 0.67 (SD 0.29) while tissue-of-origin features achieved AUC 0.77 (SD 0.26), outperforming NfL (AUC 0.53) and GFAP (AUC 0.59) in historical comparisons. As we plan for validation in follow-up studies with contemporaneous benchmarking, these preliminary findings suggest that cfDNA methylation and its inferred tissue-of-origin as promising biomarkers for MS diagnosis and subtype classification.

### cfDNA methylation level and its inferred tissue-of-origin distinguished disability severity

We evaluated whether cfDNA methylation profiles and tissue-of-origin patterns could distinguish disability severity levels at sample collection for disability monitoring. Using a well-validated and already clinically implemented patient-reported outcome (PRO), Patient Determined Disease Steps (PDDS), we categorized pwMS as having severe (PDDS≥4) vs non-severe (*i.e.,* normal-mild-moderate, PDDS<4) disability status based on the need for ambulatory assistance around blood collection. (This clinically available PRO is a more convenient outcome than other rater-assessed disability outcomes not collected in current clinical practice.) We identified 521 hypermethylated and 923 hypomethylated CpGs in the severe disability group compared to the non-severe disability group (q value<0.1 and absolute methylation difference>0.1) (**Fig. 3A, Supplementary Table 7**). DMRs, merged from adjacent differentially methylated CpGs, clearly distinguished these two disability groups (**Fig. 3B, Supplementary Table 8, Supplementary Fig. 1B**). Notably, these DMRs were highly enriched in active promoter and TSS-flanking regions and depleted in the quiescent regions (**Fig. 3C**), suggesting strong gene-regulatory potential of these DMRs. Gene Ontology analysis identified significant enrichment of cell adhesion, nervous system development, and anatomical structure development (**Fig. 3D, Supplementary Fig. 6, Supplementary Table 9)**. Motif analysis revealed substantial overrepresentation of multiple transcription factor binding sites, including members of the AP-2 family previously implicated in cranial neural crest cell development^52^ (**Supplementary Fig. 7, Supplementary Table 9**). Tissue-of-origin pattern analysis showed lower monocyte cell death and significant variability in epithelial cell death in the severe disability group (**Fig. 3E, Supplementary Fig. 8, Supplementary Table 6**, one-side Mann-Whitney U test, p<0.05). These observations corroborated previous studies linking peripheral blood monocyte counts to MS severity^53^ while supporting future studies on the potential role for epithelial cells in MS progression.^54^

**Figure 3.**
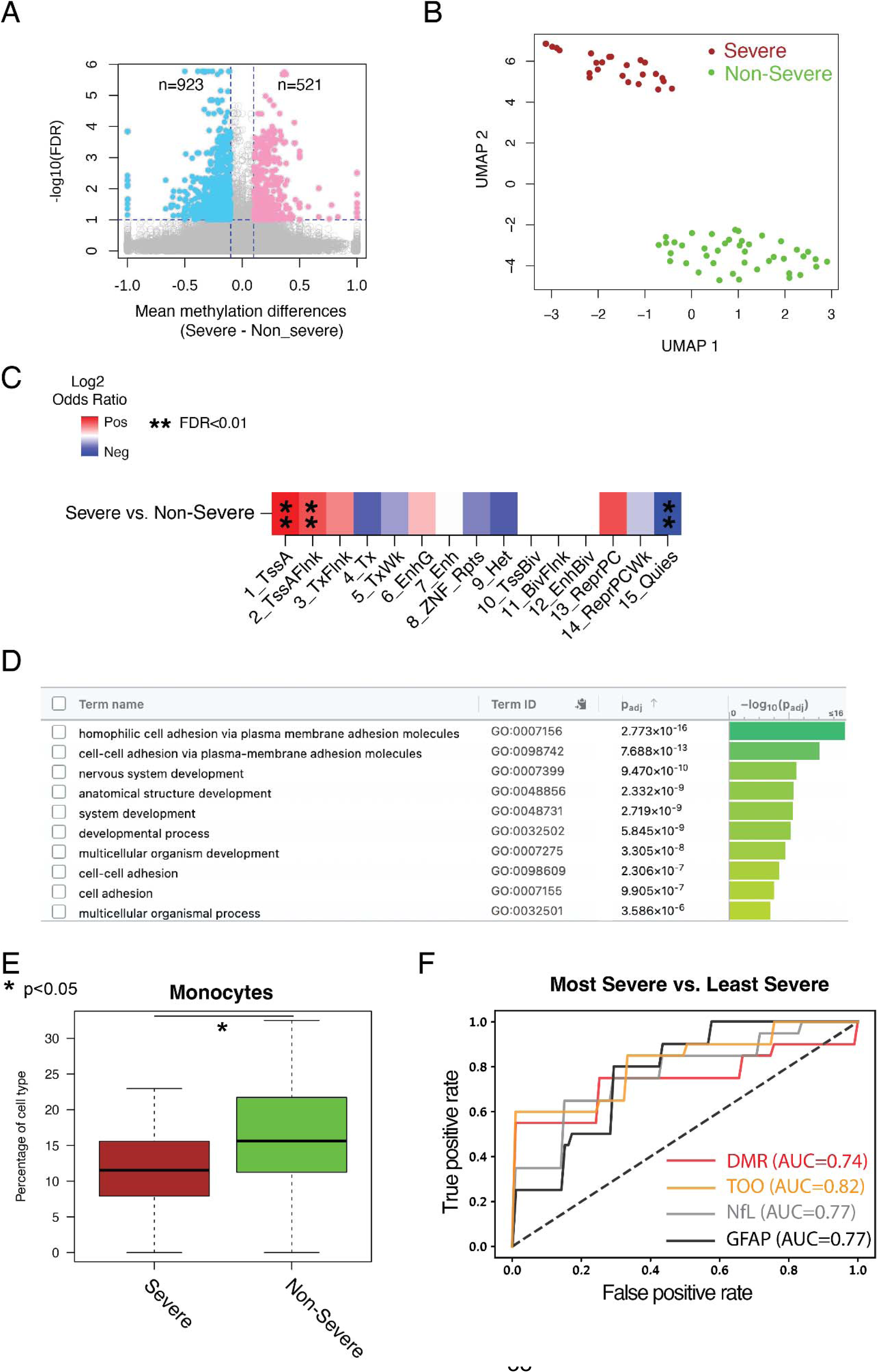
Circulating cfDNA methylation and its inferred tissue-of-origin patterns distinguished individuals with different disability levels. **A**. Scatter plots of -log10(FDR) vs mean methylation differences at differentially methylated CpGs between two disability severity levels based on the requirement for ambulatory assistance around sample collection: higher PDDS scores (severe disability, PDDS≥4) vs lower PDDS scores (non-severe, *i.e.,* normal-mild-moderate disability, PDDS<4). Colors indicated CpGs with FDR<0.1 and absolute mean methylation difference>0.1. PDDS score (*i.e.,* patient determined disease steps) indicated patient-reported ambulatory disability status. **B**. Uniform Manifold Approximation and Projection (UMAP) representation of cfDNA methylation level in DMRs could distinguish pwMS based on the binary threshold of patient-reported disability (PDDS≥4 vs <4). **C**. Enrichment and depletion of DMRs in different chromHMM states (that were previously computed from GM12878 cells in the Epigenome Roadmap project) when comparing between pwMS with severe disability (PDDS≥4) vs non-severe disability (PDDS<4). **D**. Gene ontology enrichment analysis of DMRs between pwMS with severe disability (PDDS≥4) vs non-severe disability (PDDS<4) with prioritization by g:profile. **E**. Differences in cell death contribution from Monocytes when comparing between pwMS with severe disability (PDDS≥4) vs non-severe disability (PDDS<4). (See **Supplementary Figure 8** for various Epithelial cell types.) **F**. Receiver operating characteristic (ROC) curve by DMR only (red), Tissue-of-origin only (TOO, orange), NfL only (grey), and GFAP only (black) to distinguish MS patients based on patient-reported disability: Most Severe Disability (PDDS≥6) vs Least Severe (*i.e.,* Normal-Mild, PDDS≤1). NfL and GFAP were measured from a subset of the participants from the same cohort as approximate historical benchmarks.

We explored the predictive performance of cfDNA methylation profiles in distinguishing pwMS with the most severe disability (PDDS≥6) from those with the least severe (*i.e.,* normal-mild) disability (PDDS≤1) using 10-fold cross-validation. First, we identified DMRs and differentially inferred tissue-of-origin patterns (in immune and epithelial cells) in each training fold. Then, we applied machine learning models for evaluation in the corresponding held-out test fold. cfDNA methylation DMRs alone achieved AUC 0.74 (SD 0.34), while tissue-of-origin patterns alone yielded AUC 0.82 (SD 0.23) (**Fig. 3F)**. Both approaches performed comparably to historical comparisons with NfL and GFAP (AUC 0.77 each) in a subset of the same cohort, suggesting that cfDNA methylation and its inferred tissue-of-origin had the *potential* to predict MS disease severity at blood collection, though further confirmation is required given the less robust predictive performance when attempting to predict PDDS≥4 vs <4 using the current dataset (with or without confounder adjustment), possibly due to the modest sample size limiting statistical power (data not shown).

### cfDNA methylation level at prognostic regions could predict future disability progression

Finally, we explored whether baseline cfDNA methylation profiles could predict future disability progression. Using baseline cfDNA methylation levels and longitudinal PDDS scores (**Supplementary Table 10**), we fitted a linear mixed-effects model to assess the interaction between baseline methylation and time. After adjusting for covariates (age, sex, race, DMT type and effectiveness, disease duration, and follow-up time) and correcting for multiple tests, we identified 5,392 “prognostic regions” (FDR<0.01, bonferroni correction) where their baseline methylation levels were significantly associated with the disability severity trajectory over time (**Fig. 4A**; **Supplementary Fig. 9**; **Supplementary Table 11**). To evaluate the predictive performance, we deployed a gradient boosting model via cross-validation, using baseline methylation at these prognostic regions as features and PDDS scores measured approximately 1,000 days post–blood draw as the outcome. After repeating the procedure 100 times, the model differentiated pwMS with the most severe disability (PDDS≥6) from those with the least (*i.e.,* normal-mild) disability (PDDS≤1) (AUC 0.74, SD 0.16; **Fig. 4B**).

**Figure 4.**
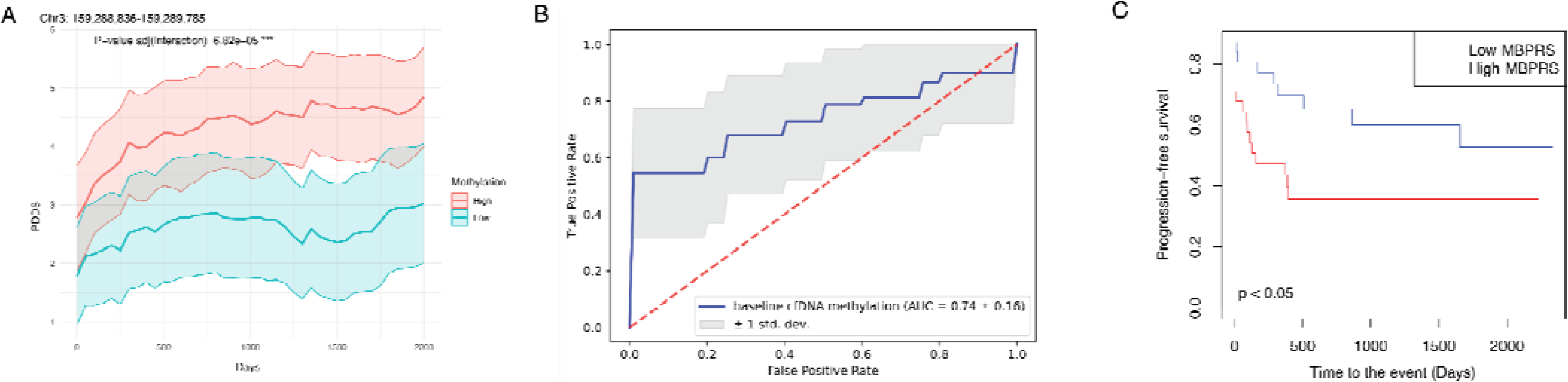
Baseline circulating DNA methylation profiles informed future disability severity trajectory. **A**. Representative plots for the patient-reported disability severity (*i.e.,* PDDS) trajectories over time stratified according to baseline circulating cfDNA methylation level (high, ≥median, red; low, <median, green) at one representative prognostic genomic region. The group with higher methylation levels showed increasing trajectory in patient-reported disability severity over time. The p-value was corrected for multiple testing (Bonferroni). The shaded area represented the 95% confidence interval. *PDDS*, patient determined disease steps. **B**. Receiver operating characteristic (ROC) curve using the baseline cfDNA methylation level at 5,392 prognostic regions to predict the binary status of patient-reported disability severity at 1,000 days after the blood sample collection: the most severe disability (PDDS≥6) vs the least (*i.e.,* normal-mild) disability (PDDS≤1). **C**. Progression-free survival curve using the risk score calculated from the weighted sum of baseline circulating cfDNA methylation level at the top 100 most informative prognostic regions. The prognostic regions were categorized based on the product of absolute value of effect size (time × biomarker interaction term) and -log 10 FDR (time × biomarker interaction term) in each region. The p-value was calculated by the log-rank test. MBPRS score (high, ≥median, red; low, <median, blue) stratified patients into significantly different disability progression trajectories (*i.e.,* higher MBPRS predicted *worse* progression-free survival). *MBPRS*, Methylation-based progression risk score.

To begin translating these findings into a clinically actionable metric, we computed a cfDNA methylation–based progression risk score (MBPRS) for each patient using baseline methylation levels at the top 100 (most informative) prognostic regions (see details in Methods). MBPRS stratified patients into significantly different disability progression trajectories (log-rank test p<0.05; **Fig. 4C**). Short of an independent validation cohort, we performed two internal control analyses. First, MBPRS using the bottom 100 (least informative) prognostic regions yielded substantially lower discrimination (log-rank test p=0.2; **Supplementary Fig. 10A**). Second, MBPRS using the same effect sizes from these top regions but with cfDNA methylation level from 100 randomly selected, matched control regions (matched chromosomes and region sizes) over 1,000 permutations confirmed that the observed separation was highly unlikely to occur by chance (permutation p<0.05, **Supplementary Fig. 10B**). Taken together, these findings supported future clinical validation of baseline cfDNA methylation level at prognostic regions in forecasting long-term disability progression.

## Discussions

In this proof-of-concept study to assess the potential clinical utility of cfDNA methylation profiles as MS biomarkers for multiple clinical contexts, we generated low-coverage (∼1.5×) WGBS data from 75 plasma cfDNA samples, including MS subtypes and non-MS controls. Using a beta-binomial regression model, we identified thousands of differentially methylated CpGs and hundreds of DMRs despite challenges due to low-coverage sequencing. cfDNA methylation levels at these DMRs and their inferred tissue-of-origin patterns distinguished pwMS from controls, differentiated across MS subtypes, and correlated with disability severity. We further identified putative gene-regulatory functions for these DMRs and distinct tissue-of-origin patterns among participant groups. Baseline cfDNA methylation profiles showed promise in predicting future disability status within a 3-year evaluation window. Finally, a baseline summary score (*i.e.,* MBPRS) stratified patients into groups with differential disability progression trajectories.

This study has several strengths and novelties. First, this was the first whole-genome investigation of plasma cfDNA methylation and its tissue-of-origin pattern as biomarkers in MS clinical contexts to our knowledge. The tissue-of-origin patterns among MS subtypes and pwMS with differential disability severity not only confirmed previous knowledge but also highlighted the less-appreciated roles of other peripheral immune cell types in MS pathogenesis for future validation. Second, this study demonstrated the potential of circulating cfDNA methylation profile and its tissue-of-origin pattern as biomarkers to *concurrently* diagnose MS, classify MS subtype, evaluate disability severity status, and predict long-term disability progression trajectory. Our preliminary evidence suggested that circulating cfDNA methylation profile as a single assay for MS diagnosis, subtyping, and monitoring and prognosis could potentially complement established blood biomarkers such as NfL and GFAP. Third, this study leveraged samples collected from a well-characterized real-world prospective clinic cohort with a disability outcome measure (*i.e.,* PDDS) already deployed longitudinally in routine clinical care. The novelty lies in the use of longitudinal clinical outcome data to identify prognostic regions in baseline cfDNA methylation profile and to potentially inform individualized clinical guidance and eventually improve patient outcomes.

As a proof-of-concept study, the study limitations had crucially informed future study design. First, existing clinical and demographic profiles in this pilot with a modest sample size might have restricted potential interpretations. For instance, we were unable to assess whether PPMS and SPMS had distinct cfDNA methylation patterns since we grouped PPMS and SPMS under PMS given the overall lower proportion of PMS than RRMS in the clinic cohort. Further, the current cfDNA methylation profile could only predict two extremes of the disability severity status (≥6 vs ≤1) but could not robustly predict more subtle severity status using an alternative PDDS threshold (≥4 vs <4), possibly attributable to the smaller phenotypic differences given the potential confounders. We are already collecting samples and data for a larger follow-up study from two independent cohorts.

Second, we initially chose low-coverage WGBS as an affordable experimental method. While cost-effective for future large-scale clinical implementation, low-coverage WGBS likely introduced substantial noise, reduced the power to detect DMRs, and impeded robust estimation of cfDNA tissue-of-origin. As CNS-derived cfDNA would constitute a relatively small fraction of the total circulating cfDNA, this limitation was relevant for MS. Simulation analyses suggested that effective coverage of at least 8× (∼13× raw coverage) could more reliably estimate cfDNA tissue-of-origin (**Supplementary Fig. 5**). We have begun generating higher coverage WGBS data in a follow-up study for better methylation signature identification and targeted methylation assay development for eventual clinical application.

Third, comparator blood biomarkers (*e.g.,* NfL, GFAP) already available from the same clinic cohort were convenient data as *approximate* benchmarks. Given that these comparisons were not always performed using identical subsets of participants and samples, we interpreted benchmark comparisons with caution. We have begun assaying plasma NfL and GFAP in parallel to plasma cfDNA samples to prepare for future comparison.

In conclusion, this proof-of-concept study highlighted the potential for circulating cfDNA methylation profiles to serve as concurrent MS biomarkers for multiple clinical contexts. We are already planning larger-scale follow-up studies with optimized WGBS sequencing coverage (while still cost-effective) to validate these promising preliminary findings and more definitively establish the clinical utilities. Future studies will additionally incorporate multimodal data (*e.g.,* better-established blood protein biomarkers as well as standard clinical and demographic features) beyond cfDNA methylation and tissue-of-origin in multi-class classifiers to further enhance predictive performance and clinical applicability. Finally, circulating cfDNA methylation-based studies generate hypotheses for future investigation using purified cells or disease-relevant tissues (*e.g.*, CNS and peripheral immune cells) to better elucidate the underlying molecular mechanisms: *e.g.,* to disentangle whether the observed methylation differences reflect cell type-specific methylation changes or shifts in the proportion of cells contributing to cfDNA.

## Materials and Methods

### Ethics approval

The University of Pittsburgh Institutional Review Board approved the study (STUDY19080007) in accordance with the Declaration of Helsinki. All participants provided written informed consent to participate in the research.

### Clinic cohort and sample collection

Participants enrolled in a *clinic*-based long-term natural history cohort study (Prospective Investigation of Multiple Sclerosis in the Three Rivers Region [PROMOTE], Pittsburgh) donated samples during 2017-2021. The current study (**Fig. 1**) included 57 samples from adults (≥18 years of age) with a neurologist-confirmed diagnosis of MS according to the 2017 McDonald criteria and 18 samples from adults without MS as controls. Clinical and demographic data were obtained from electronic health records and the cohort registry. MS subtype and relapse status were determined by the treating neurologists. An acute relapse could be a clinical and/or radiologic event within 3 months of sample collection. We defined clinical relapses as having new or recurrent neurological symptoms (deemed MS-relevant by clinicians) lasting persistently ≥24 hours without fever or infection (≥30 days from the onset of a preceding event). We defined a radiological relapse as having either a new T1-enhancing lesion and/or a new or enlarging T2-fluid-attenuated inversion recovery (FLAIR) hyperintense lesion based on clinical radiology reports of routine brain, orbit, or spinal cord magnetic resonance imaging studies. Participants completed patient-reported outcomes (PROs) that have already been clinically implemented in local practice, including Patient Determined Disease Steps (PDDS), via electronic or paper questionnaires. Research venous blood samples were collected during clinic visits. Plasma samples were isolated within four hours of phlebotomy by centrifugation first at 2,000g for 10 minutes, and then at 15,000g for 10 minutes at room temperature, and stored at -80°C until cfDNA extraction and sequencing.

### Whole-genome bisulfite sequencing of cfDNA

cfDNA was extracted from 2mL plasma using the MagMAX™ Cell-Free DNA Isolation Kit (Applied Biosystems A29319) following the manufacturer’s protocol. The concentration and size distribution of cfDNA were measured using Qubit 1X dsDNA High Sensitivity kit (Invitrogen) and BioAnalyzer (Agilent). 5ng cfDNA underwent bisulfite conversion by the EZ-96 DNA Methylation-Gold Kit (Zymo D5008). Subsequently, we prepared cfDNA methylation libraries using the Accel-NGS® Methyl-Seq DNA Library Kit (Swift 30096) and Methyl-Seq Unique Dual Indexing Kit (SWIFT 390384). To minimize batch effects, we processed all cfDNA samples in a single batch for both bisulfite conversion and library preparation. Qualified libraries were pooled and sequenced on the Illumina NovaSeq S6000 PE150 platform with a 20% spike of PhiX.

### Pre-process of whole-genome bisulfite sequencing data

We pre-processed paired-end WGBS from cfDNA using an internal pipeline. Based on FastQC results on the distribution of four nucleotides along the sequencing cycle, the adapter was trimmed by Trim Galore! (v0.6.0)^55^ with cutadapt (v2.1.0)^56^ and with parameters “--clip_R1 8 --clip_R2 15 -- three_prime_clip_R1 5 --three_prime_clip_R2 5”. After the adapter trimming, reads were aligned to the human genome (GRCh37, human_g1k_v37.fa) by Biscuit (v0.3.16.20200420) with default parameters.^57^ PCR-duplicate reads were marked by samblaster.^58^ Only high-quality reads were used for downstream analyses (*i.e.,* uniquely mapped, properly paired, mapping quality score of 30 or greater, and not a PCR duplicate). The methylation level at each CpG was called by Bis-SNP (v0.90)^59^ using default parameters in bissnp_easy_usage.pl. Only CpGs at autosomes were used for downstream analyses.

### Identification of differentially methylated CpGs and differentially methylated regions from WGBS data

We used a beta-binomial model implemented in RADMeth^38^ to calculate DMRs between MS and controls, between different MS subtypes, and between pwMS with high and low PDDS scores, adjusting for sex as a covariate. Command *adjust* used “-bins 1:100:1” as parameters. To merge adjacent differentially methylated CpGs, we used *dmrs* with -p 0.1. The threshold to filter DMRs were q value<0.1 and absolute methylation difference>0.1.

### Visualization of differentially methylated regions from cfDNA whole-genome bisulfite sequencing data

We merged DMRs from the 75 study samples into a single location file and calculated the DNA methylation levels at these DMRs from all samples. Only DMRs with <80% missing data were retained for the analysis. We imputed the missing data using the *impute.knn* function in the *impute* package (v1.70.0) in R (v4.2.0) with parameters of “maxp="p", rowmax = 0.8”. After *scale* transformation, we applied the *svd* function in R to the methylation matrix. We plotted the first 50 singular values against the proportion of variance explained and chose the elbow point. Based on the absolute and proportion of variance explained by the top SVDs (**Supplementary Figure 1**), we chose the top six singular value decomposition components (SVDs) for DMRs between different MS subtypes, and the top four SVDs for DMRs when comparing between pwMS with high vs low PDDS scores. Then, we used *umap* (v0.2.10.0) in R for dimension reduction and visualization.

### Enrichment analysis of differentially methylated regions

We used the 15-state chromHMM result at GM12878 from the Epigenome Roadmap to compute the enrichment or depletion of DMRs. The p-value was calculated against random intervals with the same chromosome and sizes using the Fisher exact test after multiple test correction (*via* Benjamini-Hochberg). DMRs were lifted up to the hg38 genome by liftOver.^60^ Gene Ontology (GO) and motif enrichment were computed by g:profile.^61^

### Tissue-of-origin analysis using cfDNA whole-genome bisulfite sequencing data

Only high-quality reads mapped to the cell type-specific marker regions were used for the tissue-of-origin analysis. After downloading cell type-specific DNA methylation markers (Atlas.U25.l4.hg19.tsv, https://raw.githubusercontent.com/nloyfer/UXM_deconv/main/supplemental/), we converted the bam files to the pat format using *wgbstools (v0.2.2) bam2pat* with default parameters and estimated the tissue-of-origin fraction in each sample using *uxm deconv* with default parameters.^35^

### Whole-genome bisulfite sequencing coverage simulation for assessing tissue-of-origin accuracy

We merged the processed bam files from non-MS control samples, filtered low-quality reads, and achieved ∼20× effective coverage representing the population average. We randomly sampled from the merged bam files at different genomic coverages ranging from 0.01× to 12× and estimated tissue-of-origin fraction as mentioned above at each step. We repeated this process 100 times to build a distribution and estimated variance around the sample mean at different coverages.

### Linear mixed-effects model for examining the interaction effect between baseline cfDNA methylation level and time on disability severity trajectory

To evaluate the association between baseline biomarker (cfDNA methylation) levels and disability progression over time, we conducted a linear mixed-effects analysis using the *lmer* function from the *lme4* package (v1.1)^62^ in R (v4.2.0). The outcome variable was repeated measures of patient-reported disability severity (*e.g.*, PDDS scores), collected at multiple time points for each patient after sample collection for baseline cfDNA methylation profiling. Fixed effects included: (1) time (as a continuous variable), (2) baseline biomarker group (categorized as “High” vs “Low” based on median cfDNA methylation levels in specific genomic regions), and (3) an interaction term (time × biomarker group) to assess potential differences in disability severity trajectories between groups. We controlled for age (continuous), sex, race, disease duration, DMT type at sample collection, DMT effectiveness at sample collection, and follow-up duration (time from sample collection to outcome assessment) as covariates. (This study population included only one ethnicity group, *i.e.,* non-Hispanic). A random intercept for each participant accounted for repeated within-person observations.

The model specification is as follows:

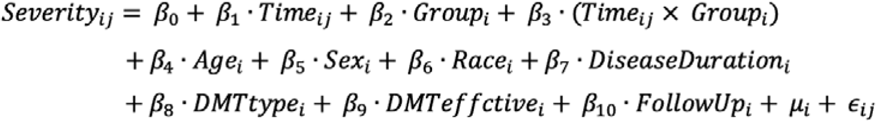

Where:

- Severity_ij_: Disability severity for patient i at time j.
- β_0_: Fixed intercept.
- β_1_: Fixed effect of time.
- β_2_: Fixed effect of biomarker group (cfDNA methylation level).
- β_3_: Interaction effect between time and biomarker group.
- β_4_, β_5_, and β_6_ represent the effects of age, sex, and race, respectively.
- β_7_: Disease duration time.
- β_8_: DMT type at sample collection.
- β_9_: DMT effectiveness at sample collection.
- β_10_: Follow-up duration time.
- u_i_: Random intercept for patient i.
- □_ij_: Residual error term.

We used the *lmerTest* package (v3.1)^63^ to obtain p-values for the fixed effects using Satterthwaite’s method for approximating degrees of freedom. We checked model assumptions by examining residual plots for homoscedasticity and normality, set statistical significance at a two-sided alpha level of 0.01, and corrected for multiple comparisons using the Bonferroni procedure when testing multiple biomarkers to control the false discovery rate.

### PDDS interpolation

Given the pragmatic study design of the *clinic*-based cohort, PDDS scores were not always collected at identical time intervals for all participants. To address this, we performed linear interpolation to facilitate the analysis of baseline cfDNA methylation profiles in relation to longitudinal trajectories of PDDS scores. Using the *interp1d* function from *SciPy* (v1.12.0),^64^ we interpolated each participant’s scores at 50-day intervals, covering the period from day 0 to day 2,000 after blood sample collection.

### Candidate genomic regions for the linear mixed model

We first selected genomic regions without missing data between pwMS with higher PDDS (PDDS≥4) and lower PDDS score (PDDS<4) at baseline. We then merged regions located within 300 bp of one another using bedtools,^65^ resulting in 2,316,371 total regions. We excluded samples containing >90% missing values across these regions in subsequent analyses.

### Methylation-based prognostic risk score and progression-free survival analysis

We began by selecting prognostic regions from the linear mixed-effects model at a false discovery rate of <0.01. We ranked regions by the product of their absolute value of estimated effect size (time × biomarker interaction term) and −log₁₀(FDR) (time × biomarker interaction term) and retained the top 100 informative regions. For each participant, we calculated the methylation-based prognostic risk score (MBPRS) as the weighted sum of baseline cfDNA methylation at these regions, using the corresponding effect sizes (time × biomarker interaction term) as weights. Missing methylation values were imputed via the impute.knn function (impute v1.70.0) in R (v4.2.0) with default settings.

Progression-free survival (PFS) was analyzed using the survival package (v3.8) in R (v4.2.0). The event time was defined as the first visit with PDDS≥4 (*i.e.,* requiring ambulatory assistance). We dichotomized patients into “High” vs “Low” MBPRS groups based on the median score of baseline cfDNA methylation level of the prognostic regions and assessed group differences in PFS by the log-rank test.

To validate MBPRS specificity, we generated 1,000 sets of matched random genomic intervals (matched chromosomes and region sizes). For each set, we extracted baseline methylation levels, applied the original weights to compute the control MBPRS scores, and performed the same log-rank test. From the resulting distribution of permutation p-values, we derived an empirical p-value for our observed separation.

### Machine learning models: general comments

For all machine learning methods described below, we evaluated seven commonly used machine learning models and reported the best performing model: LogisticRegression via “l2” or “elasticnet”, support vector machine (SVM) via “rbf” or “linear” kernels, XGBoost, GradientBoostingClassifier, and LightGBM via LGBMClassifier. Parameter tuning was conducted within each model. For DMR features, we tested various transformations (*e.g.*, raw input, top 2–20 SVD components, recursive feature elimination). As there is currently no unified model or an ensemble approach, these explorations were justified.

### Machine learning models for comparing MS vs non-MS controls

We employed a 10-fold cross-validation scheme. In each fold, 90% of the MS cases and 90% of the controls constituted the training set, while the remaining 10% served as the held-out test set. This approach ensured balanced case vs control ratios in both training and test sets. Using a beta-binomial model (RADMeth), we identified differentially methylated CpGs and merged adjacent sites to form DMRs (used *dmrs* in RadMeth with -p 0.1). Methylation densities were computed for each DMR. Missing data were marked as NA and processed using NumPy’s nan_to_num function. We then performed singular value decomposition (SVD) and selected the top two SVD components as input features based on the proportion of variance explained. We trained a logistic regression model (l2 LogisticRegression from scikit-learn, v1.1.1)^66^ with the following parameters:

“penalty=’l2’, dual=False, tol=1e-5, C=120, fit_intercept=True, intercept_scaling=1, class_weight=’balanced’, solver=’lbfgs’, max_iter=1000”.

To calculate all possible differential tissue-of-origin patterns in CNS and immune cells, we conducted a one-sided Mann-Whitney U test to identify cell types (within CNS and immune cell types) that differed significantly (p<0.05) in group comparisons. Cell types meeting this criterion in any comparison were included as features. We trained a gradient boosting classifier (GradientBoostingClassifier from scikit-learn) with the following parameters:

“n_estimators=200, learning_rate=0.2, max_depth=10, min_samples_split=2, min_samples_leaf=1, subsample=0.8, max_features=’log2’”.

Because each participant had only one historical blood NfL or GFAP value, we applied a simple logistic regression model (C=1.0, using *StratifiedKFold* for the 10-fold cross-validation).

### Machine learning models for comparing PMS vs RRMS

We used the same 10-fold cross-validation scheme and preprocessing steps (as in the MS vs controls comparison). For DMRs features, instead of taking the top SVD components, we employed recursive feature elimination with cross-validation (RFECV from scikit-learn) in each fold to refine the final features (“step=1,scoring=’accuracy’,min_features_to_select=1”). We trained a LightGBM classifier (LGBMClassifier in *lightgbm*) with the following parameters:

“n_estimators=100, learning_rate=0.1, num_leaves=31, max_depth=1, reg_alpha=0.1, reg_lambda=0.1, subsample=0.8, colsample_bytree=0.8, min_child_samples=10”.

For differential tissue-of-origin features, given the smaller sample size of PMS relative to RRMS, we relaxed the tissue-of-origin p-value cutoff to 0.1. We again performed a one-sided Mann-Whitney U test (p<0.1) to select significantly altered cell types. We trained a SVM with the following parameters: “C=8.0, kernel=’rbf’, probability=True”.

### Machine learning models for comparing actively relapsing RRMS vs stable remission RRMS

For DMR features, we retained the top three SVD components (based on the proportion of variance explained) of DMR methylation levels as input. We trained an SVM model with the following parameters: “kernel=’rbf’, probability=True, gamma=’scale’, C=0.1”.

For differential tissue-of-origin features, given the smaller sample size of actively relapsing RRMS relative to stable remission RRMS, we relaxed the p-value cutoff to 0.2. We then used a logistic regression model with the following parameters: “penalty=’l2’, dual=False, tol=0.0001, C=100, fit_intercept=True, intercept_scaling=1, class_weight=’balanced’, solver=’lbfgs’, max_iter=100”.

### Machine learning models to distinguish different disability severity levels

We excluded samples lacking PDDS scores, retaining 67. For exploring given the greater phenotypic distinction, we only used samples with PDDS≤1 (least severe, *i.e.,* mild or normal) or ≥6 (most severe), thereby reducing the sample size to 46. We applied the same 10-fold cross-validation and preprocessing strategies (as in MS vs control comparisons). For DMRs features, we used XGBoost^67^ with the following parameters:

“max_depth=3, learning_rate=0.4, n_estimators=1000, class_weight=’balanced’, objective=’binary:logistic’, subsample=0.667”.

For differential tissue-of-origin features, we tested all immune and epithelial cell types using a p-value cutoff of 0.1, and trained a GradientBoostingClassifier with the following parameters:

“n_estimators=200, learning_rate=0.15, max_depth=2, min_samples_split=2, min_samples_leaf=2, subsample=0.8, max_features=’log2’”.

### Machine learning approach to predict the future disease severity

We included only samples with PDDS score measurements at ≥3 different time points. We focused on the 5,392 potentially “prognostic regions” identified by the linear mixed-effects model. At ∼1,000 days post-blood draw, samples with PDDS ≤1 or ≥6 were retained for analysis. We used a RepeatedStratifiedKFold (five folds, repeated 100 times). The GradientBoostingClassifier was trained with the following parameters:

“n_estimators=100, learning_rate=0.1, max_depth=5, min_samples_split=2, min_samples_leaf=1, subsample=0.8, max_features=’log2’”.

### Statistics and Reproducibility

No statistical method was used to predetermine the sample size in this proof-of-concept pilot study. The experiments were randomized to generate cfDNA sequencing libraries. Blinding to allocation during experiments and outcome assessment were not applicable.

## Data Availability

The raw WGBS sequencing data generated in this study have been deposited in the European Genome-phenome Archive (EGA) with controlled access under accession code (pending). Data access can be obtained through a request to the corresponding authors. The corresponding authors will generally respond to requests within one week. Once granted, the access has no time restriction The raw sequencing data are protected by data privacy laws. Processed DNA methylation level is available at Gene Expression Omnibus (GEO) with access ID (pending). The processed and de-identified data are available at zenodo.org (doi: https://doi.org/10.5281/zenodo.14803482). The remaining data are available within the Article, Supplementary Information, and Source Data file.

## Code Availability

Analysis scripts are publicly available on GitHub under the MIT license for academic researchers: https://github.com/epifluidlab/ms_cfnda_wgbs_manuscript. The zipped code and meta-data are also available on zenodo.org (doi: https://doi.org/10.5281/zenodo.14803482).

## Acknowledgments

This research was supported in part through the computational resources and staff contributions provided for the Quest high performance computing facility at Northwestern University, which is jointly supported by the Office of the Provost, the Office for Research, and Northwestern University Information Technology. This research was also supported in part through the computational resources and staff contributions provided by the Genomics Compute Cluster, which is jointly supported by the Feinberg School of Medicine, the Center for Genetic Medicine, and Feinberg’s Department of Biochemistry and Molecular Genetics, the Office of the Provost, the Office for Research, and Northwestern Information Technology. The Genomics Compute Cluster is part of Quest, Northwestern University’s high performance computing facility, with the purpose to advance research in genomics. This work also used the Extreme Science and Engineering Discovery Environment (XSEDE), which is supported by the National Science Foundation grant number ACI-1548562. This work used the XSEDE at the Pittsburgh Supercomputing Center (PSC) through allocation MCB190124P and MCB190006P. L.W. and Y.L. are supported by the startup grant to Y.L. from Cincinnati Children’s Hospital Medical Center, Northwestern University, Robert H. Lurie Comprehensive Cancer Center of Northwestern University, and the iDEA-TECH award from Sanofi Inc. (to Y.L. and Z.X.). Z.X. is supported by NINDS (R01NS098023 and R01NS124882).

## Author Contributions

L.W., Y.L., and Z.X. conceived the study. H.F. and L.W. performed the cfDNA extraction and library constructions. Y.L., R.B., and K.H. performed the data analysis with input from Z.X.. Z.X., W.Z., and L.Z. provided the clinical samples and guidance related to the clinic applications. H.F., R.B., W.Z., L.W., Y.L., and Z.X. wrote the manuscript together. All authors read and approved the final manuscript.

