## Supplementary Tables for "Circulating cell-free DNA methylation profiles as noninvasive multiple sclerosis biomarkers: A proof-of-concept study"

### Supplementary Figures

#### Supplementary Figure 1. Variance explained by the top singular value decompositions (SVDs).

**A.** Absolute (left) and proportion (right) of variance explained by the top SVDs for the cfDNA methylation level from differentially methylated regions (DMRs) that were called across MS subtypes. **B.** Absolute (left) and proportion (right) of variance explained by the top SVDs for the cfDNA methylation level from DMRs that were called between the two disability severity levels based on the requirement of ambulatory assistance around sample collection: higher PDDS scores (severe disability, PDDS≥4) and lower PDDS scores (non-severe, *i.e.,* normal-mild-moderate disability, PDDS<4). *PDDS*, patient determined disease steps.

A

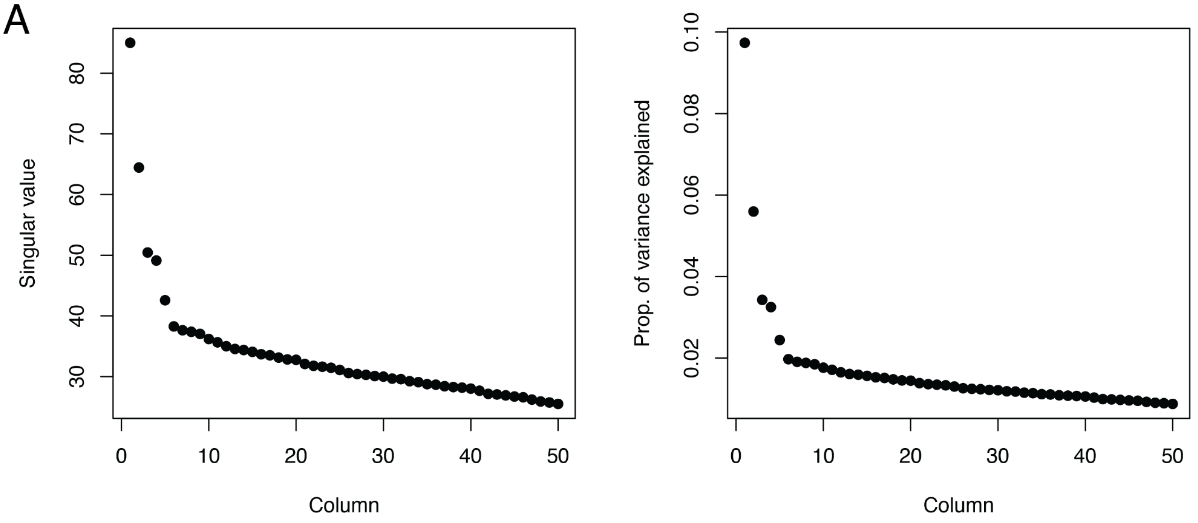


B

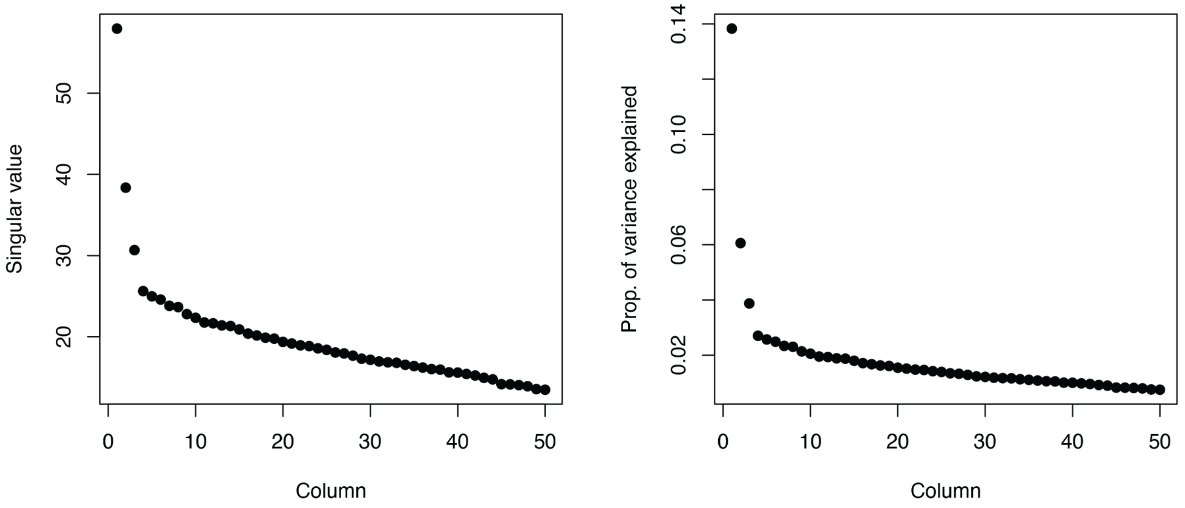

**Supplementary Figure 2. Gene ontology analysis on differentially methylated regions (DMRs).**

**A.** PMS vs RRMS. **B.** A-RRMS vs S-RRMS. *PMS*, progressive MS; *RRMS*, relapsing-remitting MS; *A-RRMS*, actively relapsing RRMS; S-RRMS, stable remission RRMS.

A

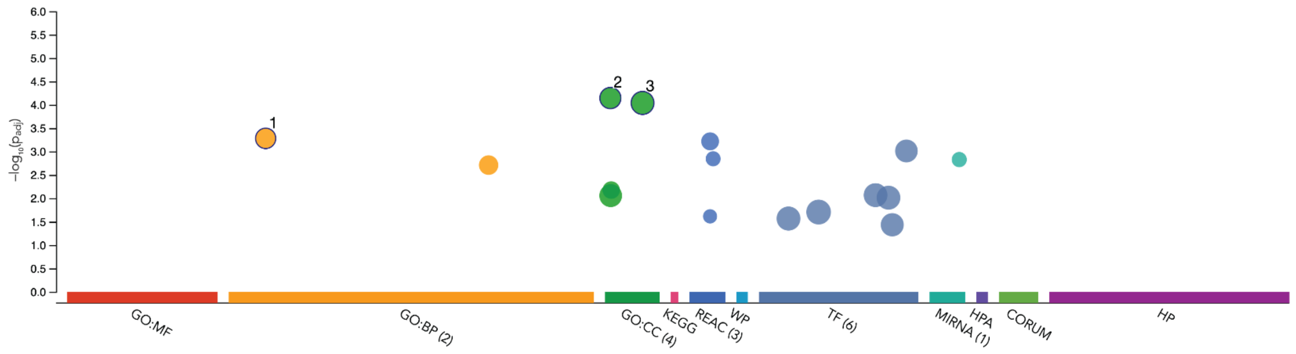


| ID | Source | Term ID | Term Name | $p_{adj}$ (query_... |
|---|---|---|---|---|
| 1 | GO:BP | GO:0007155 | cell adhesion | $5.228\times10^{-4}$ |
| 2 | GO:CC | GO:0005856 | cytoskeleton | $7.175\times10^{-5}$ |
| 3 | GO:CC | GO:0071944 | cell periphery | $9.125\times10^{-5}$ |

B

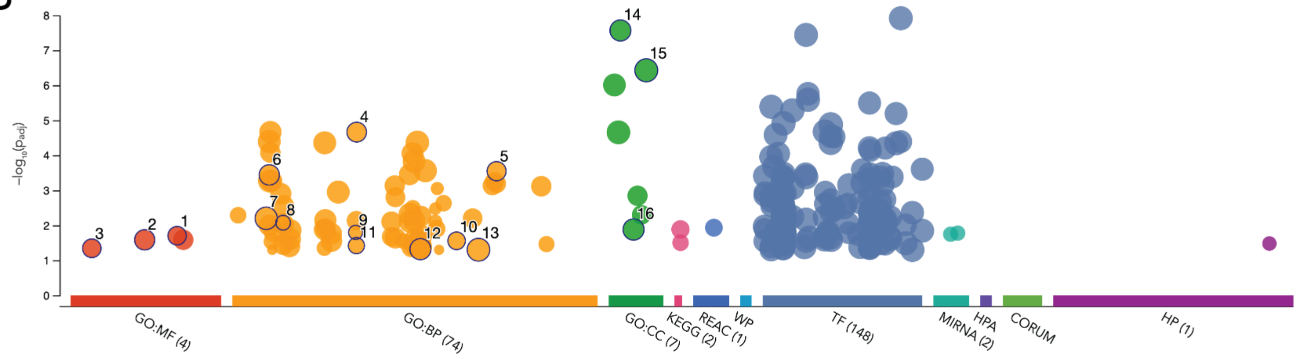


| ID | Source | Term ID | Term Name | $p_{adj}$ (query_... |
|---|---|---|---|---|
| 1 | GO:MF | GO:0050839 | cell adhesion molecule binding | $1.975\times10^{-2}$ |
| 2 | GO:MF | GO:0038023 | signaling receptor activity | $2.592\times10^{-2}$ |
| 3 | GO:MF | GO:0008134 | transcription factor binding | $4.515\times10^{-2}$ |
| 4 | GO:BP | GO:0035295 | tube development | $2.174\times10^{-5}$ |
| 5 | GO:BP | GO:0099536 | synaptic signaling | $2.833\times10^{-4}$ |
| 6 | GO:BP | GO:0007155 | cell adhesion | $3.621\times10^{-4}$ |
| 7 | GO:BP | GO:0006810 | transport | $6.285\times10^{-3}$ |
| 8 | GO:BP | GO:0009880 | embryonic pattern specification | $8.415\times10^{-3}$ |
| 9 | GO:BP | GO:0035176 | social behavior | $1.589\times10^{-2}$ |
| 10 | GO:BP | GO:0070588 | calcium ion transmembrane transport | $2.766\times10^{-2}$ |
| 11 | GO:BP | GO:0035265 | organ growth | $3.787\times10^{-2}$ |
| 12 | GO:BP | GO:0051172 | negative regulation of nitrogen compound met... | $4.794\times10^{-2}$ |
| 13 | GO:BP | GO:0080090 | regulation of primary metabolic process | $4.973\times10^{-2}$ |
| 14 | GO:CC | GO:0030054 | cell junction | $2.721\times10^{-8}$ |
| 15 | GO:CC | GO:0071944 | cell periphery | $3.738\times10^{-7}$ |
| 16 | GO:CC | GO:0042995 | cell projection | $1.316\times10^{-2}$ |

**Supplementary Figure 3. Motif enrichment at differentially methylated regions (DMRs).**

**A.** MS vs controls. **B.** PMS vs RRMS. **C.** A-RRMS vs S-RRMS. Enrichment was calculated by g:profile. *PMS*, progressive MS; *RRMS*, relapsing-remitting MS; *A-RRMS*, actively relapsing RRMS; S-RRMS, stable remission RRMS.

A

| Term name | Term ID | $p_{adj}$ |
|---|---|---|
| Factor: ZBTB14; motif: SSCCGCGCACNS | TF:M12461 | $3.845\times10^{-4}$ |
| Factor: IRX2a; motif: ACRYGNNNNACRYGT; match ... | TF:M11022_1 | $5.134\times10^{-3}$ |
| Factor: BEN; motif: CAGCGRNV; match class: 1 | TF:M01240_1 | $6.840\times10^{-3}$ |
| Factor: ZIC4; motif: NNCCNCCCRYNGYGN | TF:M12227 | $1.059\times10^{-2}$ |
| Factor: TCF-1; motif: ACATCGRGRCGCTGW; match... | TF:M11601_1 | $1.362\times10^{-2}$ |
| Factor: SATB2; motif: GSCGCTGTCCNNGGTGCTG... | TF:M12660_1 | $1.892\times10^{-2}$ |
| Factor: E2F-1:HES-7; motif: GGCRCGTGSYNNWN... | TF:M08525 | $1.938\times10^{-2}$ |
| Factor: RERE; motif: CNGCNSCNNGSRCRGSGSS | TF:M13129 | $2.030\times10^{-2}$ |
| Factor: SATB2; motif: GSCGCTGTCCNNGGTGCTGN | TF:M12660 | $2.039\times10^{-2}$ |
| Factor: ZNF138; motif: GCAGCRSCNSGSNCMGSG... | TF:M13144_1 | $2.080\times10^{-2}$ |
| Factor: ZNF432; motif: NCAGNRCCNSRGRCAGC; ... | TF:M12704_1 | $2.095\times10^{-2}$ |
| Factor: MAZ; motif: GGGMGGGGS; match class: 1 | TF:M10432_1 | $2.505\times10^{-2}$ |
| Factor: BEN; motif: CAGCGRNV | TF:M01240 | $3.182\times10^{-2}$ |
| Factor: ZNF219; motif: SNNCAGCACCNNGGNCAG... | TF:M13149_1 | $3.210\times10^{-2}$ |
| Factor: ZNF142; motif: SNSCGCCCGCCCGCSCSS;... | TF:M12697_1 | $3.693\times10^{-2}$ |
| Factor: Egr-1; motif: NGCGTGCGY; match class: 1 | TF:M04950_1 | $4.259\times10^{-2}$ |

B

| Term name | Term ID | $p_{adj}$ |
|---|---|---|
| Factor: ZNF609; motif: GNSNGGGNGCTGN; match... | TF:M13132_1 | $9.720\times10^{-4}$ |
| Factor: TFAP2A; motif: NGCCCYNNGGGCN; match... | TF:M04148_1 | $8.564\times10^{-3}$ |
| Factor: ZNF138; motif: GCAGCRSCNSGSNCMGSG... | TF:M13144 | $9.699\times10^{-3}$ |
| Factor: IRX-1; motif: NACRYNNNNNNNNRYGNN; m... | TF:M11018_1 | $1.968\times10^{-2}$ |
| Factor: ETF; motif: GVGGMGG | TF:M00695 | $2.712\times10^{-2}$ |
| Factor: ZNF253; motif: SNGNSCGNGGNGCKGNN; ... | TF:M13147_1 | $3.683\times10^{-2}$ |

C

| Term name | Term ID | $p_{adj}$ |
|---|---|---|
| Factor: ZNF37A; motif: CCYYGGCTCCNTSCCMN | TF:M12354 | $1.215\times10^{-8}$ |
| Factor: GKLF; motif: NNNRGGNGNGGSN; match cl... | TF:M07289_1 | $3.647\times10^{-8}$ |
| Factor: HA95; motif: CCSNSSCCNSCNCCWSCCNS... | TF:M13127_1 | $1.748\times10^{-6}$ |
| Factor: HA95; motif: CCSNSSCCNSCNCCWSCCNS | TF:M13127 | $2.588\times10^{-6}$ |
| Factor: Sp1; motif: RGGGMGGRGSNGGGG | TF:M10529 | $3.223\times10^{-6}$ |
| Factor: BTEB3; motif: CCNNSCCNSCCCCKCCCCC | TF:M09826 | $4.115\times10^{-6}$ |
| Factor: ETF; motif: CCCCGCCCCYN; match class: 1 | TF:M07039_1 | $5.260\times10^{-6}$ |
| Factor: ZNF253; motif: SNGNSCGNGGNGCKGNN; ... | TF:M13147_1 | $6.433\times10^{-6}$ |
| Factor: E2F-4; motif: SNGGGCGGGAANN; match c... | TF:M09894_1 | $1.224\times10^{-5}$ |
| Factor: MAZ; motif: GGGMGGGGSSGGGGGGGG... | TF:M09636 | $1.324\times10^{-5}$ |

**Supplementary Figure 4. Tissue-of-origin estimated from different MS subtypes and non-MS controls.**

One-side Mann-Whitney U test was performed between every two groups of participants (*e.g.*, PMS vs A-RRMS, or PMS vs A-RRMS+S-RRMS+Control). The side was determined if the median was larger or smaller between the groups. *PMS*, progressive MS; *RRMS*, relapsing-remitting MS; *A-RRMS*, actively relapsing RRMS; *S-RRMS*, stable remission RRMS.  *, p<0.05; **, p<0.01.

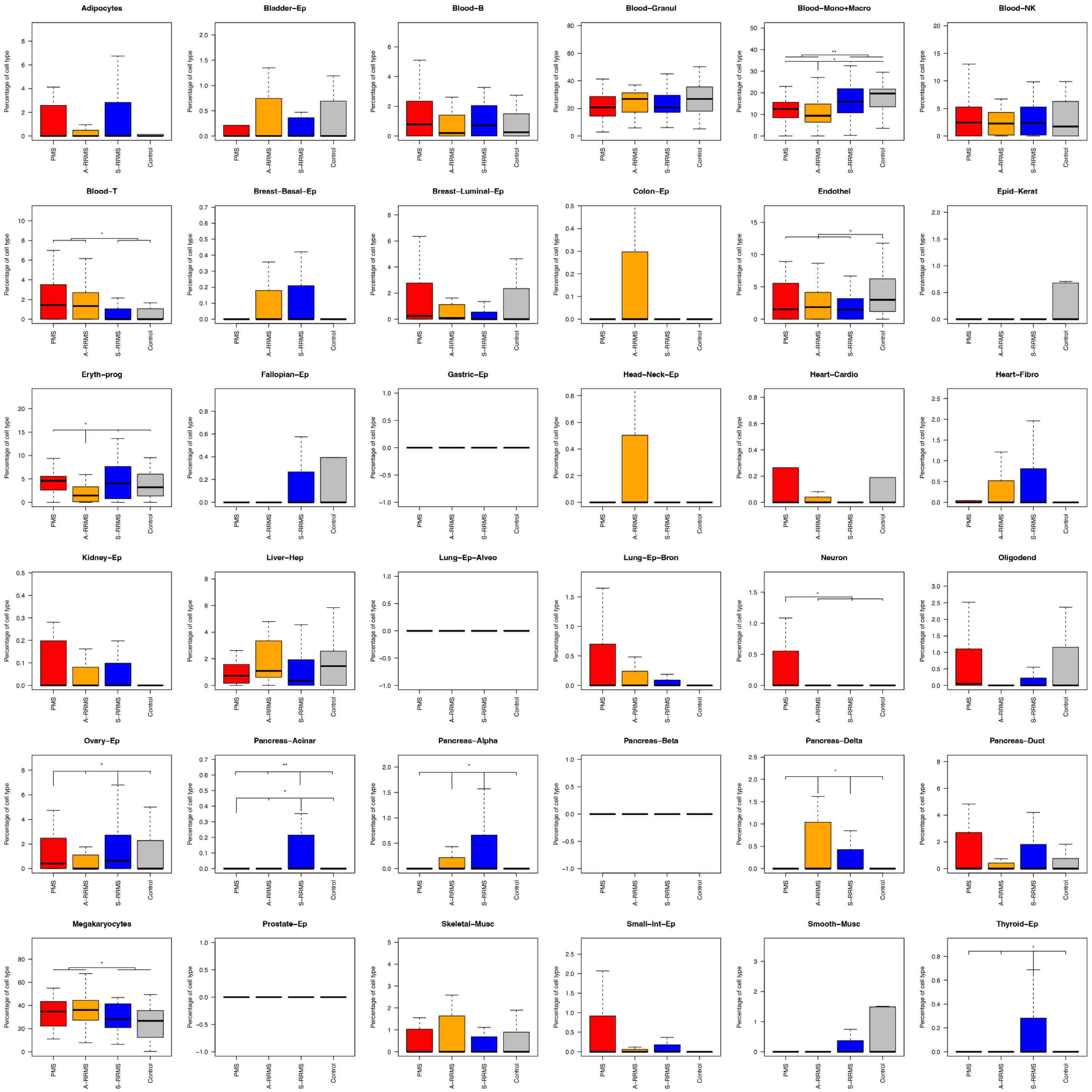

**Supplementary Figure 5. Simulation results on the variation of identifying tissue-of-origin at different coverages of cfDNA WGBS.**

**A.** Tissue fractions at different downsampled coverages across tissues and cell types. **B.** The standard deviation of tissue fraction at different downsampled coverages across tissues and cell types.

A

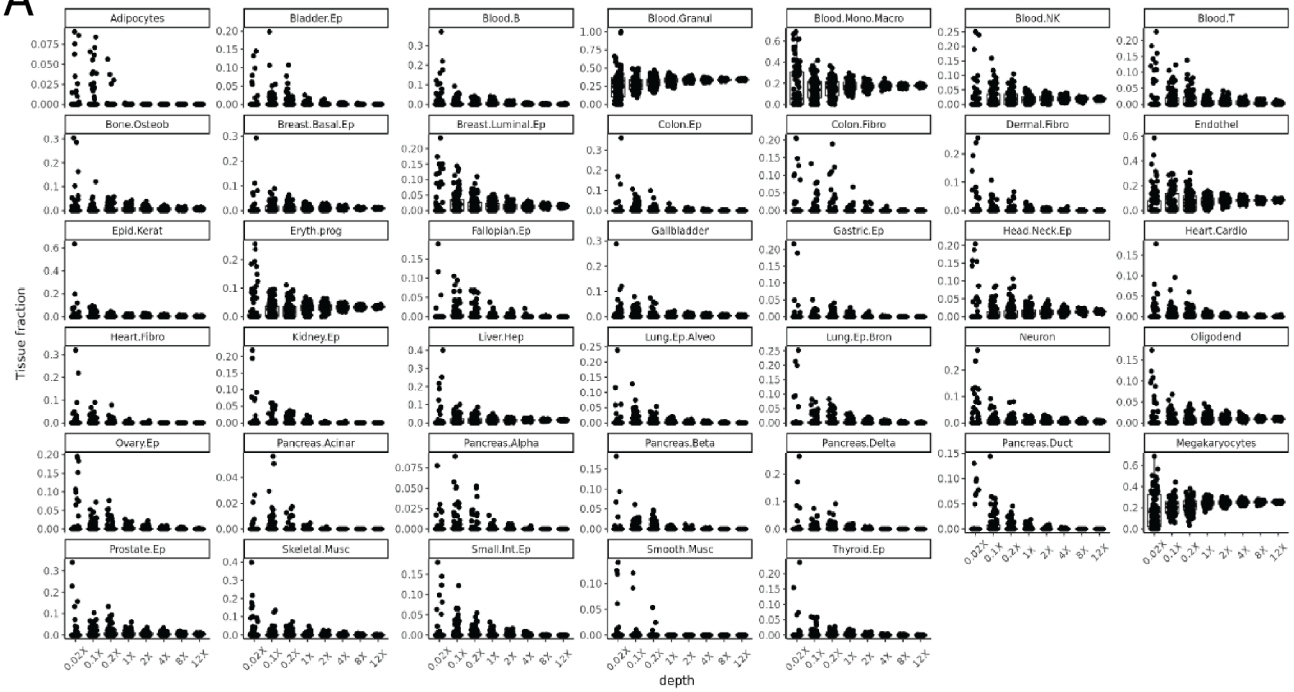


B

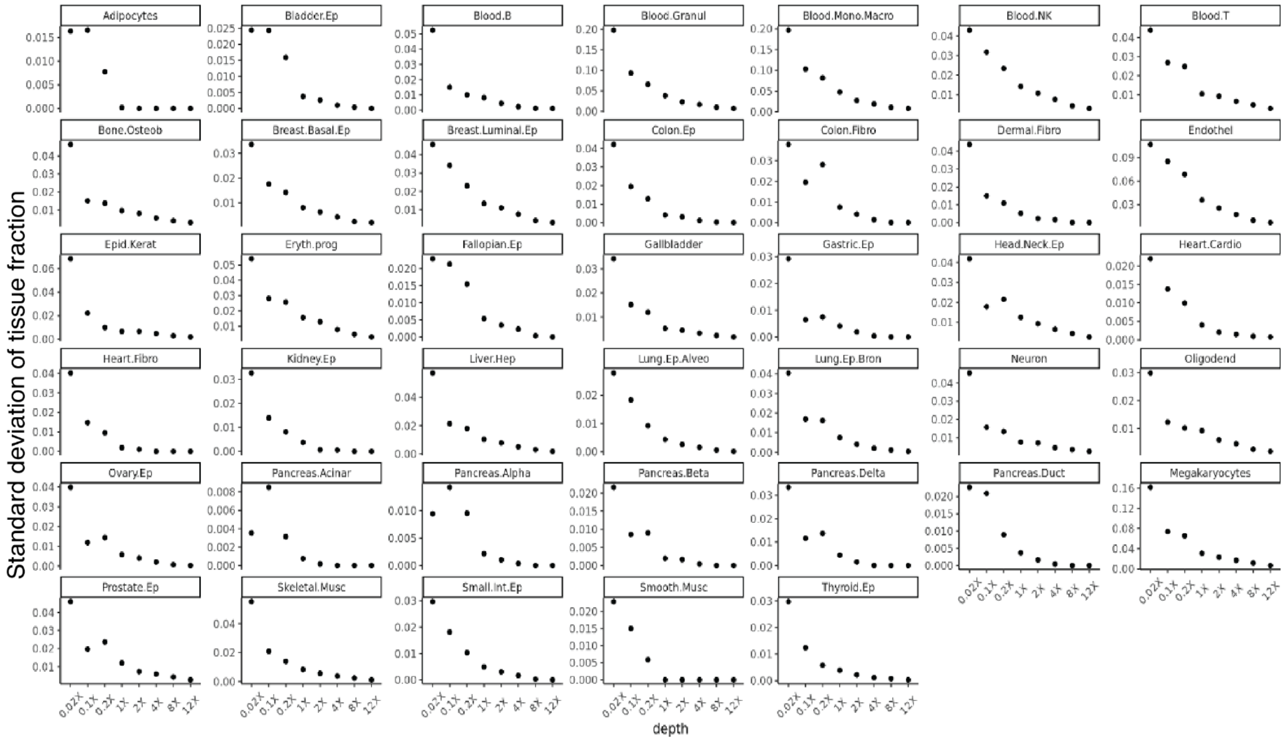

**Supplementary Figure 6. Gene ontology analysis on differentially methylated regions (DMRs) between disability severity groups.**

Two disability severity groups were categorized based on the requirement for ambulatory assistance around sample collection: higher PDDS scores (severe disability, PDDS≥4) and lower PDDS scores (non-severe, *i.e.,* normal-mild-moderate disability, PDDS<4). *PDDS*, patient determined disease steps.

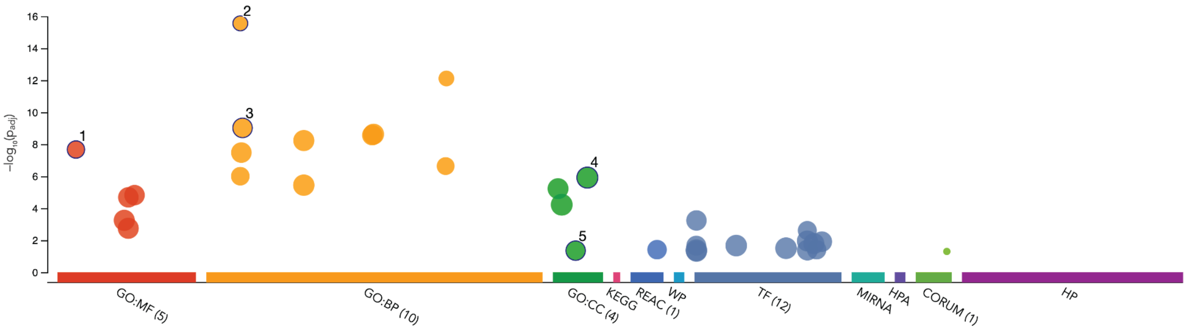


| ID | Source | Term ID | Term Name | $p_{adj}$ (query_... |
|---|---|---|---|---|
| 1 | GO:MF | GO:0005509 | calcium ion binding | $2.124 \times 10^{-8}$ |
| 2 | GO:BP | GO:0007156 | homophilic cell adhesion via plasma membrane... | $2.773 \times 10^{-16}$ |
| 3 | GO:BP | GO:0007399 | nervous system development | $9.470 \times 10^{-10}$ |
| 4 | GO:CC | GO:0071944 | cell periphery | $1.207 \times 10^{-6}$ |
| 5 | GO:CC | GO:0042995 | cell projection | $4.435 \times 10^{-2}$ |

**Supplementary Figure 7. Motif enrichment on differentially methylated regions (DMRs) between disability severity groups.**

Two disability severity groups were categorized based on the requirement for ambulatory assistance around sample collection: higher PDDS scores (severe disability, PDDS≥4) and lower PDDS scores (non-severe, *i.e.,* normal-mild-moderate disability, PDDS<4). *PDDS*, patient determined disease steps.

| Term name | Term ID | $p_{adj}$ | $-\log_{10}(p_{adj})$ 0 ≤16 |
|---|---|---|---|
| Factor: AP-2; motif: GSCCSCRGGCNRNRNN; matc... | TF:M00800_1 | $5.770\times10^{-4}$ | |
| Factor: WT1; motif: NGCGGGGGGGTSMMCYN; ma... | TF:M05327_1 | $2.435\times10^{-3}$ | |
| Factor: WT1; motif: NGCGGGGGGGTSMMCYN | TF:M05327 | $1.103\times10^{-2}$ | |
| Factor: ZNF383; motif: SSNGGGMGGNGSNGGS; ... | TF:M12703_1 | $1.217\times10^{-2}$ | |
| Factor: ZNF138; motif: GCAGCRSCNSGSNCMGSG... | TF:M13144_1 | $1.605\times10^{-2}$ | |
| Factor: HA95; motif: CCSNSSCCNSCNCCWSCCNS... | TF:M13127_1 | $2.146\times10^{-2}$ | |
| Factor: AP-2beta; motif: GCNNNGGSCNGVGGGN; ... | TF:M01858_1 | $2.168\times10^{-2}$ | |
| Factor: RNF96; motif: BCCCGCRGCC | TF:M01199 | $3.134\times10^{-2}$ | |
| Factor: ZNF232; motif: NCAGCASCNNGGNCAGCG... | TF:M13146_1 | $3.923\times10^{-2}$ | |
| Factor: WT1; motif: CGCCCCCNCN; match class: 1 | TF:M02036_1 | $4.096\times10^{-2}$ | |
| Factor: AP-2gamma; motif: GCCYNCRGSN | TF:M03811 | $4.163\times10^{-2}$ | |
| Factor: AP-2; motif: MKCCCSCNGGCG; match clas... | TF:M00189_1 | $4.849\times10^{-2}$ | |

**Supplementary Figure 8. Tissue-of-origin estimated between disability severity groups.**

Two disability severity groups were categorized based on the requirement for ambulatory assistance around sample collection: higher PDDS scores (severe disability, PDDS≥4) and lower PDDS scores (non-severe, *i.e.,* normal-mild-moderate disability, PDDS<4). One-side Mann-Whitney U test was performed between groups of patients. The side was determined if the median was larger or smaller between the groups. *PDDS*, patient determined disease steps. *, p<0.05.

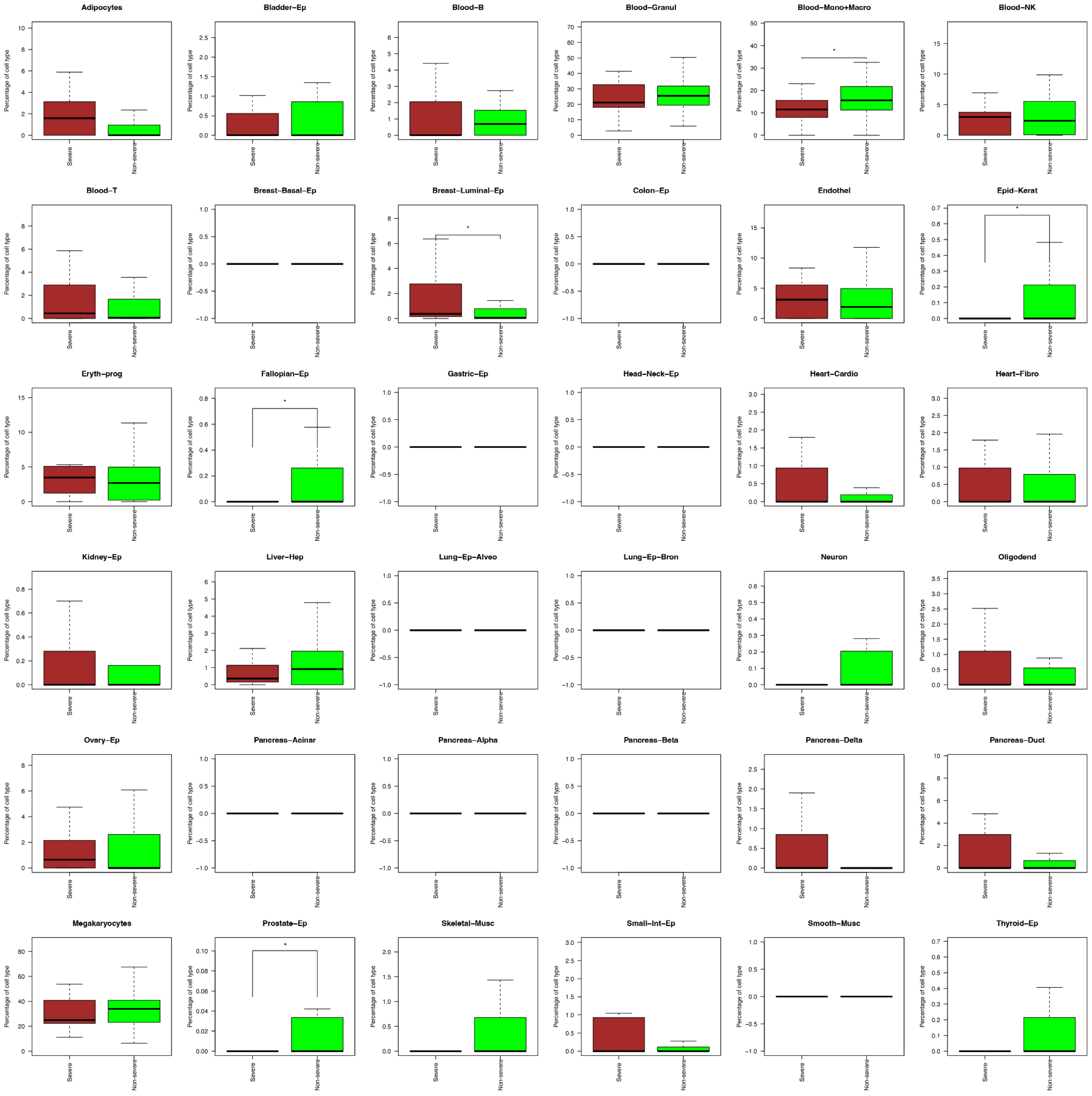

**Supplementary Figure 9. Additional representative plots for the patient-reported disability severity trajectories over time stratified according to baseline cfDNA methylation level.**

Baseline cfDNA methylation level (high, ≥median, red; vs low, <median, green) at two additional representative prognostic genomic regions. The group with lower methylation levels (left panel) and the group with higher methylation levels (right panel) showed increasing trajectory in patient-reported disability severity over time, respectively. The p-value was corrected for multiple testing (Bonferroni). The shaded area represented the 95% confidence interval. *PDDS*, patient determined disease steps.

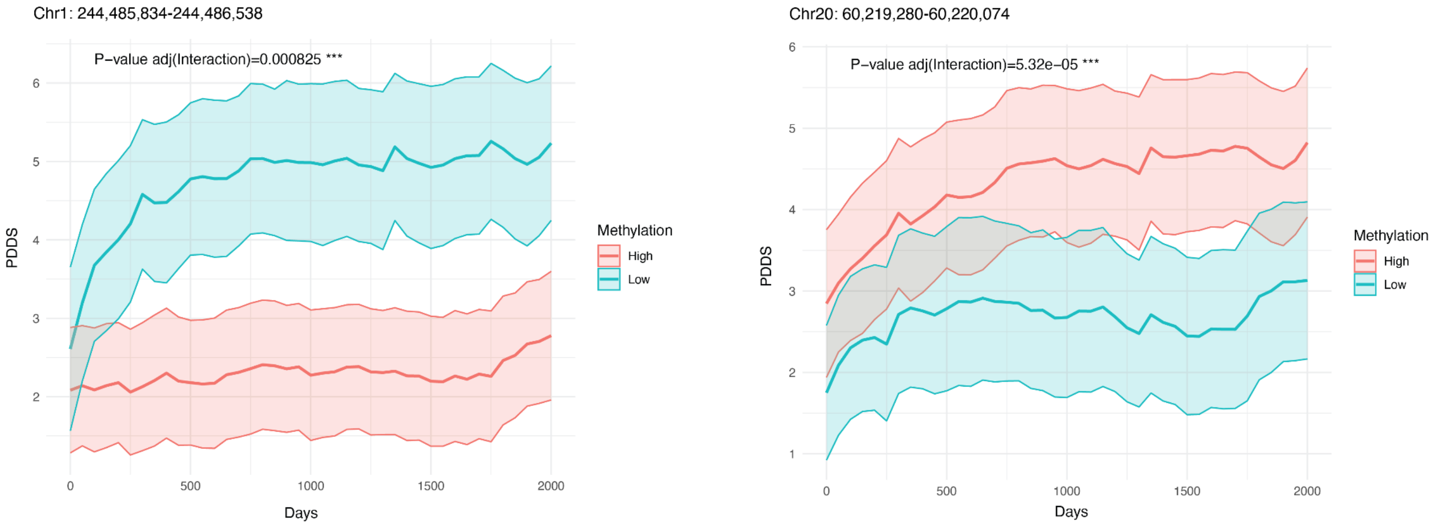

**Supplementary Figure 10. Log-rank test p value from two different sets of control regions.**

**A.** Progression-free survival curve using the risk score calculated from the weighted sum of baseline cfDNA methylation level at 100 least informative prognostic regions. The prognostic regions were categorized based on the product of absolute value of effect size (time × biomarker interaction term) and -log 10 FDR (time × biomarker interaction term) in each region. The p value was calculated by the log-rank test. This "control" MBPRS score (high, ≥median, red; low, <median, blue) did *not* reach statistical significance via log rank test despite visible separation of survival curves. *MBPRS*, Methylation based progression risk score. **B.** The distribution of permutated p-values calculated from the matched random control regions (matched chromosomes and region sizes). MBPRS for each patient were calculated using the original effect size (from the top 100 regions) and baseline cfDNA methylation level from matched random regions. For each iteration, the log-rank test p-value was calculated between the two groups of patients with a different MBPRS score. The process was repeated 1,000 times to obtain the p-value distribution. The observed log-rank test p-value from the top 100 informative prognostic regions (Figure 4C) was marked as the red line here.

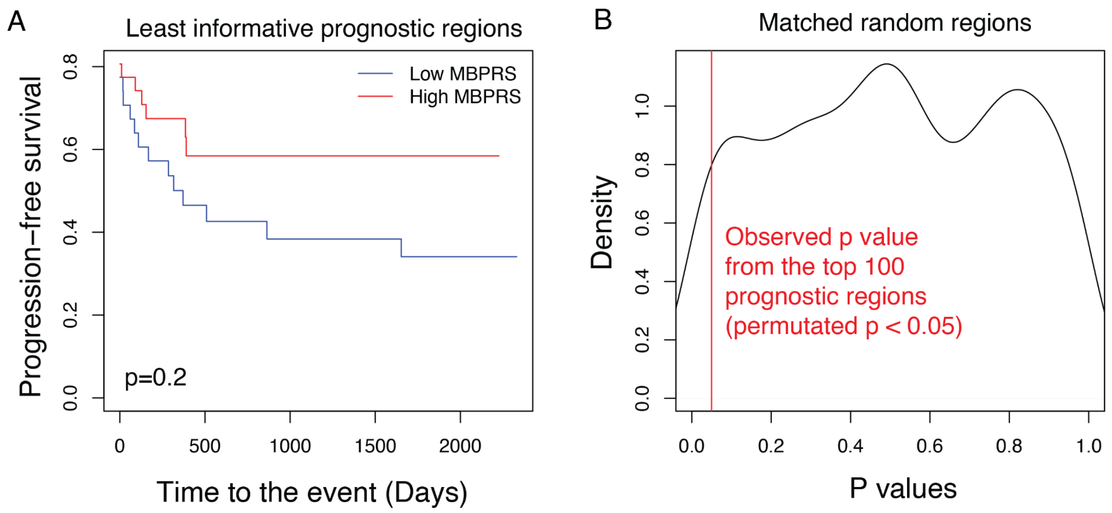

### Supplementary Tables

**Supplementary Table 1**. Participant-level demographic and clinical profile. (As this table provided individual level data, we showed age range instead of age to protect confidentiality per guideline.)

**Supplementary Table 2**. Summary statistics of cfDNA WGBS.

**Supplementary Table 3**. Differential CpG sites in cfDNA WGBS comparing MS and controls and comparing across MS subtypes.

**Supplementary Table 4**. Differentially methylated regions in cfDNA WGBS comparing MS and controls and comparing across MS subtypes.

**Supplementary Table 5**. Gene ontology and motif enrichment at differentially methylated regions in cfDNA WGBS comparing MS and controls and comparing across MS subtypes (from g:profile).

**Supplementary Table 6**. Tissue-of-origin inferences from cfDNA WGBS in each sample.

**Supplementary Table 7**. Differential CpG sites in cfDNA WGBS comparing MS patients with higher vs lower patient-reported disability (PDDS≥4 vs <4).

**Supplementary Table 8**. Differentially methylated regions in cfDNA WGBS comparing MS patients with higher vs lower patient-reported disability severity (PDDS≥4 vs <4).

**Supplementary Table 9**. Gene ontology and motif enrichment at DMRs in cfDNA WGBS comparing MS patients with higher vs lower patient-reported disability severity (PDDS≥4 vs <4) from g:profile.

**Supplementary Table 10**. Post-baseline (*i.e.,* sample collection) longitudinal patient-reported disability scores for each participant at different time points.

**Supplementary Table 11**. Prognostic genomic regions where baseline cfDNA methylation levels were associated with differential patient-reported disability progression.
